# The ongoing evolution of variants of concern and interest of SARS-CoV-2 in Brazil revealed by convergent indels in the amino (N)-terminal domain of the Spike protein

**DOI:** 10.1101/2021.03.19.21253946

**Authors:** Paola Cristina Resende, Felipe G Naveca, Roberto D. Lins, Filipe Zimmer Dezordi, Matheus V. F. Ferraz, Emerson G. Moreira, Danilo F. Coêlho, Fernando Couto Motta, Anna Carolina Dias Paixão, Luciana Appolinario, Renata Serrano Lopes, Ana Carolina da Fonseca Mendonça, Alice Sampaio Barreto da Rocha, Valdinete Nascimento, Victor Souza, George Silva, Fernanda Nascimento, Lidio Gonçalves Lima Neto, Fabiano Vieira da Silva, Irina Riediger, Maria do Carmo Debur, Anderson Brandao Leite, Tirza Mattos, Cristiano Fernandes da Costa, Felicidade Mota Pereira, Cliomar Alves dos Santos, Darcita Buerger Rovaris, Sandra Bianchini Fernandes, Adriano Abbud, Claudio Sacchi, Ricardo Khouri, André Felipe Leal Bernardes, Edson Delatorre, Tiago Gräf, Marilda Mendonça Siqueira, Gonzalo Bello, Gabriel L Wallau, on behalf of Fiocruz COVID-19 Genomic Surveillance Network

**Affiliations:** Laboratory of Respiratory Viruses and Measles (LVRS), Instituto Oswaldo Cruz, FIOCRUZ-Rio de Janeiro, Brazil; Laboratório de Ecologia de Doenças Transmissíveis na Amazônia (EDTA), Instituto Leônidas e Maria Deane, FIOCRUZ-Amazonas, Brazil; Department of Virology, Instituto Aggeu Magalhães, FIOCRUZ-Pernambuco, Brazil; Departamento de Entomologia, Instituto Aggeu Magalhães, FIOCRUZ-Pernambuco, Brazil; Núcleo de Bioinformática (NBI), Instituto Aggeu Magalhães FIOCRUZ-Pernambuco, Brazil; Department of Fundamental Chemistry, Federal University of Pernambuco, Recife, Brazil; Laboratório Central de Saúde Pública do Estado do Maranhão (LACEN-MA), Brazil; Laboratório Central de Saúde Pública do Estado do Paraná (LACEN-PR), Brazil; Laboratório Central de Saúde Pública do Estado do Alagoas (LACEN-AL), Brazil; Laboratório Central de Saúde Pública do Amazonas (LACEN-AM), Brazil; Fundação de Vigilância em Saúde do Amazonas, Brazil; Laboratório Central de Saúde Pública do Estado da Bahia (LACEN-BA), Brazil; Laboratório Central de Saúde Pública do Estado de Sergipe (LACEN-SE), Aracajú, Sergipe, Brazil; Laboratório Central de Saúde Pública do Estado de Santa Catarina (LACEN-SC), Florianópolis, Santa Catarina, Brazil; Instituto Adolfo Lutz, São Paulo, São Paulo, Brazil; Laboratório de Enfermidades Infecciosas Transmitidas por Vetores, Instituto Gonçalo Moniz, FIOCRUZ-Bahia, Salvador, Bahia, Brazil; Laboratório Central de Saúde Pública do Estado de Minas Gerais (LACEN-MG); Departamento de Biologia. Centro de Ciências Exatas, Naturais e da Saúde, Universidade Federal do Espírito Santo, Alegre, Brazil; Plataforma de Vigilância Molecular, Instituto Gonçalo Moniz, FIOCRUZ-Bahia, Brazil; Laboratório de AIDS e Imunologia Molecular, Instituto Oswaldo Cruz, FIOCRUZ-Rio de Janeiro, Brazil

**Keywords:** COVID-19, pandemics, antibody escape, coronavirus, communitary transmission

## Abstract

Mutations at both the receptor-binding domain (RBD) and the amino (N)-terminal domain (NTD) of the SARS-CoV-2 Spike (S) glycoprotein can alter its antigenicity and promote immune escape. We identified that SARS-CoV-2 lineages circulating in Brazil with mutations of concern in the RBD independently acquired convergent deletions and insertions in the NTD of the S protein, which altered the NTD antigenic-supersite and other predicted epitopes at this region. Importantly, we detected communitary transmission of four lineages bearing NTD indels: a P.1 Δ69-70 lineage (which can impact several SARS-CoV-2 diagnostic protocols), a P.1 Δ144 lineage, a P.1-like lineage carrying ins214ANRN, and the VOI N.10 derived from the B.1.1.33 lineage carrying three deletions (Δ141-144, Δ211 and Δ256-258). These findings support that the ongoing widespread transmission of SARS-CoV-2 in Brazil is generating new viral lineages that might be more resistant to antibody neutralization than parental variants of concern.

## Introduction

Recurrent deletions in the amino (N)-terminal domain (NTD) of the spike (S) glycoprotein of SARS-CoV-2 have been identified during long-term infection of immunocompromised patients ^1–4^ as well as during extended human-to-human transmission ^3^. Most of those deletions (90%) maintain the reading frame and cover four recurrent deletion regions (RDRs) within the NTD at positions 60-75 (RDR1), 139-146 (RDR2), 210-212 (RDR3), and 242-248 (RDR4) of the S protein ^3^. The RDRs that occupy defined antibody epitopes within the NTD and RDR regions might alter antigenicity ^3^. Interestingly, the RDRs overlap with four NTD Indel Regions (IR - IR-2 to IR-5) that are prone to gain or lose short nucleotide sequences during sarbecoviruses evolution both in animals and humans ^5,6^.

Since late 2020, several more transmissible variants of concern (VOCs) and also variants of interest (VOI) with convergent mutations at the receptor-binding domain (RBD) of the S protein (particularly E484K and N501Y) arose independently in humans ^7,8^. Some VOCs also displayed NTD deletions such as lineages B.1.1.7 (RDR2 Δ144), B.1.351 (RDR4 Δ242-244), and P.3 (RDR2 Δ141-143) that were initially detected in the United Kingdom, South Africa, and the Philippines, respectively ^3^. The VOCs B.1.1.7 and B.1.351 are resistant to neutralization by several anti-NTD monoclonal antibodies (mAbs) and NTD deletions at RDR2 and RDR4 are important for such phenotype ^9–14^. Thus, NTD mutations and deletions represent an important mechanism of immune evasion and accelerate SARS-CoV-2 adaptive evolution in humans.

Several SARS-CoV-2 variants with mutations in the RBD have been described in Brazil, including the VOC P.1 ^15^ and the VOIs P.2 ^16^, N.9 ^17^ and N.10 ^18^. With the exception of N.10, none of the other variants described in Brazil displayed indels in the NTD. Importantly, although the VOC P.1 displayed NTD mutations (L18F) that abrogate binding of some anti-NTD mAbs ^14^ and further showed reduced binding to RBD-directed antibodies, it is more susceptible to anti-NTD mAbs than other VOCs ^9–14,19^. In this study, we characterized the emergence of RDR variants within VOC and VOIs circulating in Brazil that were genotyped by the Fiocruz COVID-19 Genomic Surveillance Network between November 2020 and February 2021.

## Results and Discussion

Our genomic survey identified 35 SARS-CoV-2 sequences from seven different Brazilian states that harbor a variable combination of mutations in the RBD (K417T, E484K, N501Y) and indels in the NTD region of the S protein. These genomes were classified within lineages N.10 (n = 16), P.1 (n = 14), P.2 (n = 1) and B.1.1.28 (P1-like, n = 4) (**Table 1**). Seven VOC P.1 sequences displayed deletion Δ69-70 in the RDR1, three sequences (two VOC P.1 and one VOI P.2) displayed deletion Δ144 in the RDR2, two P.1 sequences showed a four amino acid deletion Δ141-144 in the RDR2, two P.1 sequences harbors a two amino acid deletion Δ189-190, and one P.1 sequence displayed a three amino acid deletion Δ242-244 in the RDR4. We also detected four B.1.1.28 P.1-like genomes bearing an ins214ANRN insertion upstream to RDR 3 and sharing six out of 10 P.1 lineage-defining mutations in the Spike protein (L18F, P26S, D138Y, K417T, E484K, N501Y) as well as P.1 lineage-defining mutations in the NSP3 (K977Q), NS3 (S253P) and N (P80R) proteins ^20^. The VOI N.10 displayed NTD indels Δ141-144 at RDR2, Δ211 at RDR3 and Δ256-258 close to RDR4 ^18^. Inspection of sequences available at EpiCoV database in the GISAID (https://www.gisaid.org/) at March 1st, 2021, retrieved three P.1 sequences from the Bahia state ^21^ and one B.1.1.28 sequence from the Amazonas state ^20^ with deletion Δ144 **(Table 1)**.

**Table 1.**
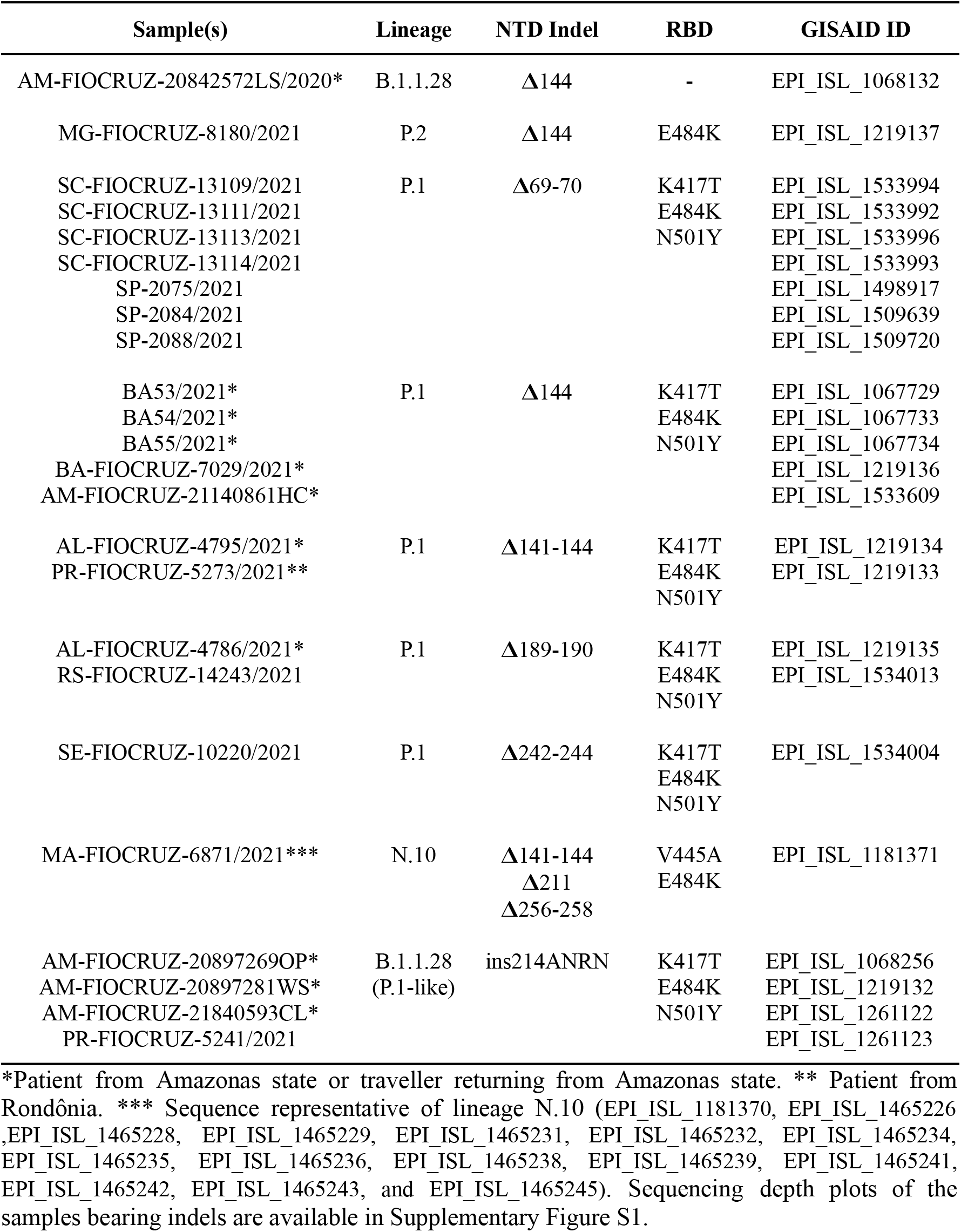
SARS-CoV-2 Brazilian variants with indels at NTD of the Spike protein.

The Maximum Likelihood (ML) phylogenetic analysis of lineage P.1 supports recurrent emergence of variants Δ141-144 and Δ69-70 and the monophyletic origin of variants Δ144 and Δ189-190 (**Fig. 1A**). Both P.1 Δ141-144 sequences recovered from patients from Amazonas and Rondônia states, all P.1 Δ69-70 sequences from Santa Catarina state and the P.1 Δ242-244 sequence from Sergipe state appeared as singletons intermixed among non-deleted P.1 sequences. The remaining P.1 variants with NTD deletions were distributed in two sub-clades that also include non-deleted P.1 sequences. One sub-clade (aLRT = 86%) was characterized by the mutations ORF1a:T951I and A18945G and comprises nine sequences: the five P.1 Δ144, the two P.1 Δ189-190 and two P.1 from Amazonas and Goiás states. The other sub-clade (aLRT = 85%) was characterized by the synonymous mutations G29781A and T29834A and comprises seven sequences: the three P.1 Δ69-70 from São Paulo state plus four P.1 sequences from São Paulo, Amazonas and Tocantins states. The ML phylogenetic analyses further confirm that all sequences belonging to variants P.1-like ins214ANRN (**Fig. 1A**) and VOI N.10 (**Fig. 1B**) branched in highly supported (aLRT > 99%) monophyletic clades. These findings revealed that NTD deletions characteristic of VOCs B.1.1.7 (Δ69-70 and Δ144) and B.1.351 (Δ242-244) occurred at multiple times during the evolution of lineage P.1 and also sporadically arose in lineages B.1.1.28, B.1.1.33 (N.10) and P.2.

**Figure 1.**
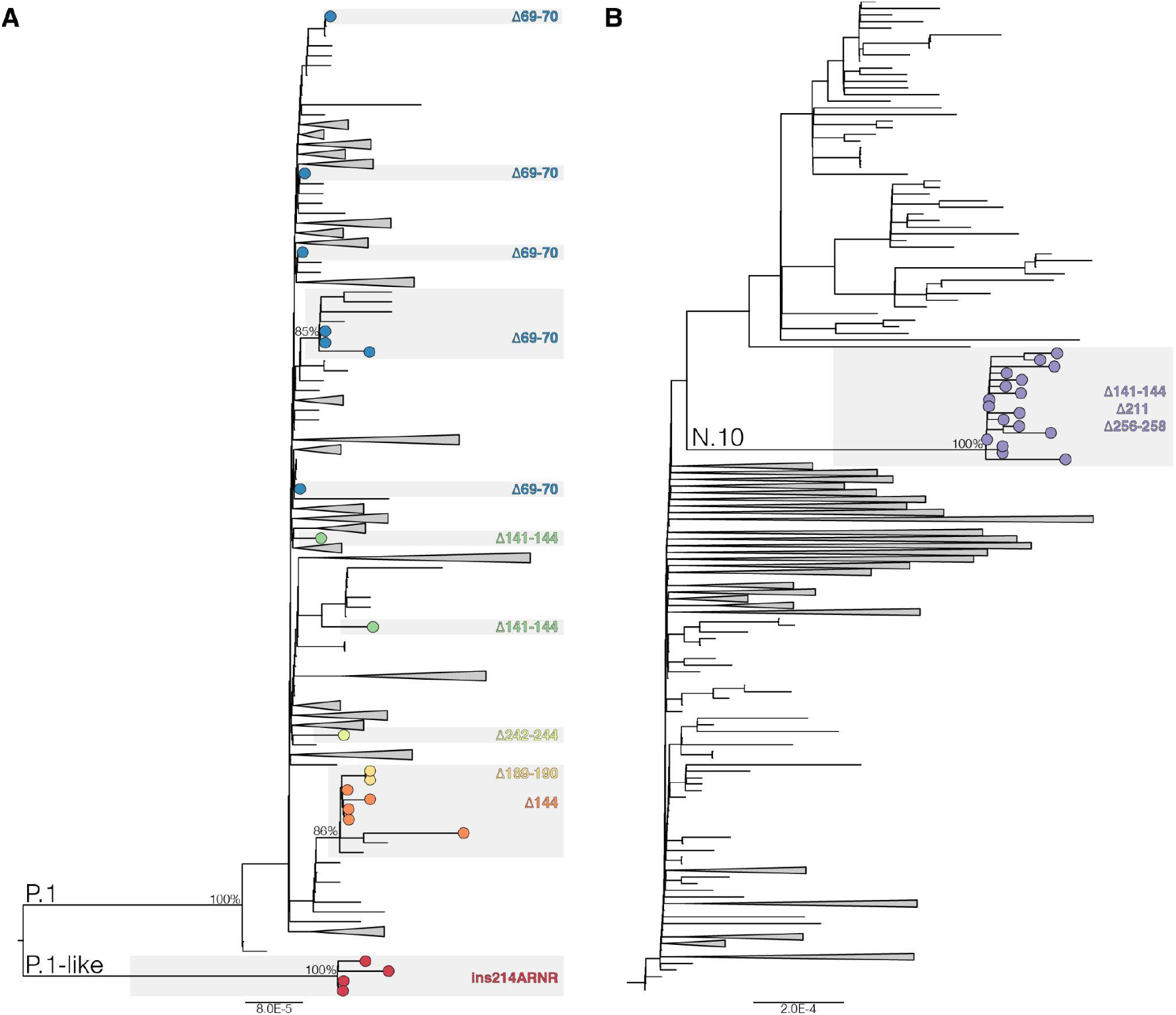
ML phylogenetic tree of whole-genome lineage P.1/P.1-like (**A**) and B.1.1.33 (**B**) Brazilian sequences showing the recurrent emergence of deletions at the NTD of the S protein. Tip circles representing the SARS-CoV-2 sequences with NTD indels are colored as indicated. The branch lengths are drawn to scale with the left bar indicating nucleotide substitutions per site. For visual clarity, some clades are collapsed into triangles.

Most P.1 Δ144, Δ141-144 and Δ189-190 sequences were detected in the Amazonas or were recovered from individuals that were transferred from or that reported a travel history to the Amazonas state, like all P.1 Δ144 sequences from the Bahia state described previously ^21^ and in the present study **(Table 1)**. Thus, those P.1 variants as well as the variant P.1-like ins214 probably emerged in the Amazonas state and some of them displayed low-level of community transmission. The P.1 Δ69-70 sequences, by contrast, were detected in autochthonous cases from Santa Catarina and Sao Paulo states that had no history of travel to the Amazonas. The phylogenetic clustering supports the independent origin of variant P.1 Δ69-70 in both Brazilian states and its local dissemination in São Paulo, but not in Santa Catarina. The lack of monophyletic clustering of P.1 Δ69-70 sequences from Santa Catarina, however, should be interpreted with caution due to the paucity of synapomorphic mutations within diversity of lineage P.1. Other variants that also arose outside the Amazonas state were the P.1 Δ242-244, the P.2 Δ144 and the VOI N.10 detected in the states of Sergipe, Minas Gerais and Maranhão, respectively ^18^.

While SARS-CoV-2 variants harboring NTD deletions at RDR2 and RDR4 have emerged in many different lineages globally, insertions in the S protein are rare events. Our search of SARS-CoV-2 sequences available at EpiCoV database in the GISAID (https://www.gisaid.org/) on March 1st retrieved only 146 SARS-CoV-2 sequences of lineages A.2.4 (n = 52), B (n = 3), B.1 (n = 7), B.1.1.7 (n = 1), B.1.177 (n = 1), B.1.2 (n = 1), B.1.214 (n = 80) and B.1.429 (n = 1) that displayed an insert motif of three to four amino acids (AKKN, KLGB, AQER, AAG, KFH, KRI, and TDR) in position 214 (**Appendix Table 1**). Most ins214 motifs were unique, except for the ins214TDR that arose independently in lineages B.1 and B.1.214. With the only exception of one lineage B sequence sampled in March 2020, all SARS-CoV-2 ins214 variants were only detected since November 2020, and its frequency increased in 2021 mainly due to the recent dissemination of lineage A.2.4 ins214AAG in Central and North America and of lineage B.1.214 ins214TDR in Europe.

To better understand the evolutionary context of NTD indels, we aligned the S protein of representative sequences of SARS-CoV-2 lineages with NTD indels and SARS-CoV-2-related coronavirus (SC2r-CoV) lineages from bats and pangolins ^22^. Inspection of the alignment confirms that most NTD indels detected in the SARS-CoV-2 lineages occur within IR previously defined in sarbecovirus (**Fig. 2**). The Δ141-144 occurs in the IR-3 located in the central part of the NTD, where some bats SC2r-CoV also have deletions. The Δ211 and ins214 occurs near the IR-4 where some bat SC2r-CoV from China (RmYN02, ins214GATP), Thailand (RacCS203, ins214GATP), and Japan (Rc-o319, ins214GATS) displayed a four amino acids insertion. Despite amino acid motifs at ins214 are very different across SARS-CoV-2 and SC2r-CoV lineages, the insertion size (3-4 amino acids) was conserved. Deletions Δ242-244 and Δ256-258 occur immediately upstream and downstream to IR-5, respectively, where some bat and pangolin SC2r-CoV lineages also displayed deletions. Thus, NTD regions that are prone to gain indels during viral transmission among animals are the same as those detected during transmissions in humans.

**Figure 2.**
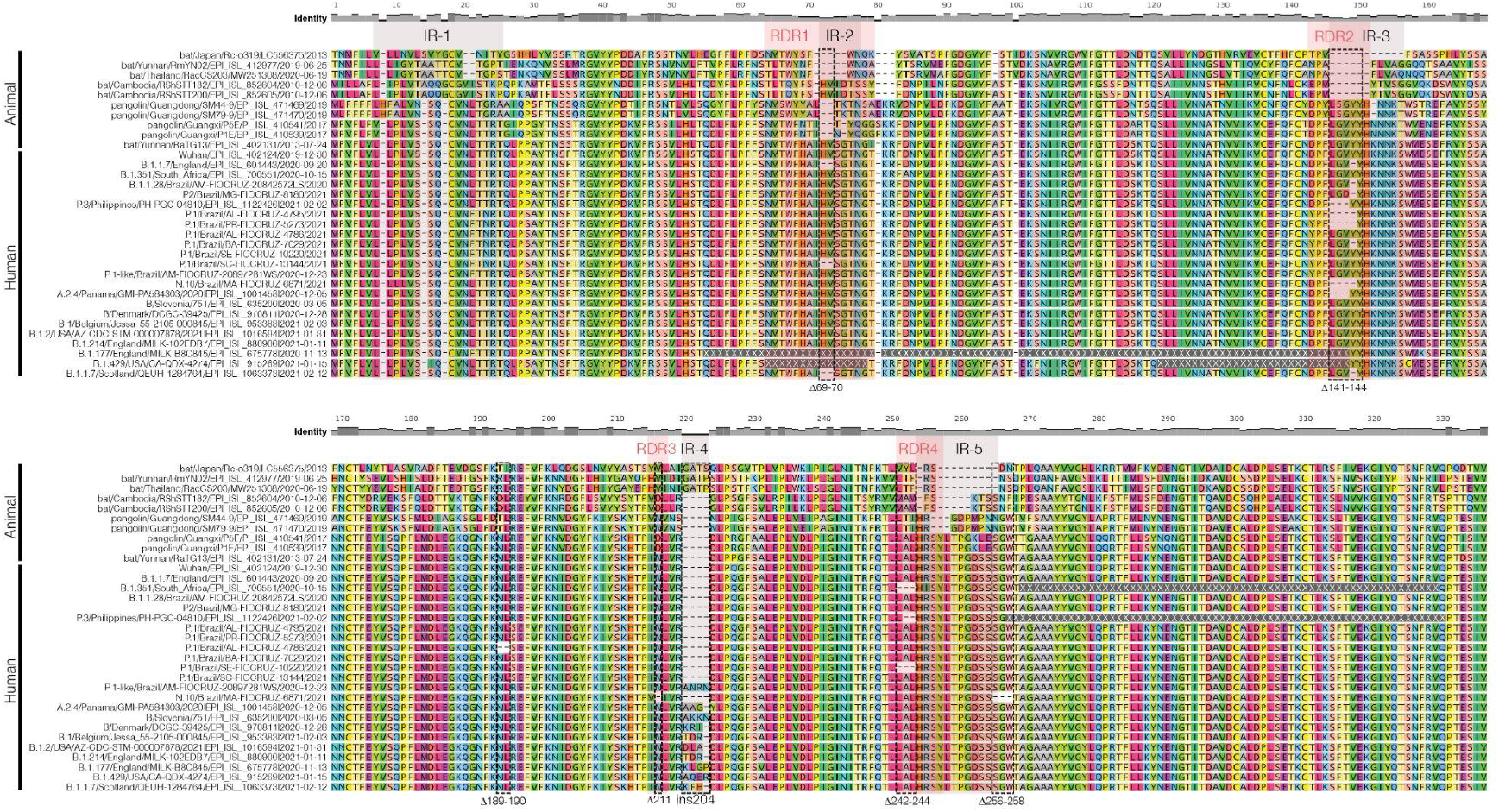
Amino acid alignment of Sarbecovirus NTD Spike region up to amino acid 335 including representative sequences of SARS-CoV-2 lineages harboring indels in the NTD and SARS-CoV-2-related coronavirus (SC2r-CoV) from bats and pangolins. IRs and RDRs positions (gray and red shaded areas, respectively) are approximations due to the high genetic variability in these alignment positions. Dotted rectangles highlight the indels identified in this study. The relative identity level estimated for each position of the alignment is displayed at the top.

Epitope mapping showed that neutralizing antibodies are primarily directed against the RBD and NTD of the S protein ^9,23–26^. Some of the RBD mutations (K417T and E484K) detected in the VOCs and VOIs circulating in Brazil have been associated with increased resistance to neutralization by mAbs, or polyclonal sera from convalescent and vaccinated subjects ^27–31^. The RDR2 and RDR4 are located in the N3 (residues 141 to 156) and N5 (residues 246 to 260) loops that composes the NTD antigenic-supersite ^32,33^ and deletions at those RDRs are also an essential mechanism for SARS-CoV-2 immune evasion of anti-NTD Abs ^3,9,10,34,14,3,9,10,34^. To further visualize the potential impact of NTD deletions on immune recognition, we performed a modeling analysis of the binding interface between the NTD region and the NTD-directed neutralizing antibody (NAb) 2-51 derived from a convalescent donor ^23,33^. The NAb 2-51 interacts with the wildtype NTD antigenic-supersite (EPI_ISL_402124) through several contacts with loops N3 and N5, with a predominance of hydrophobic contacts and dispersion interactions in N5 and saline interactions in N3 (**Fig. 3A and B**).

**Figure 3.**
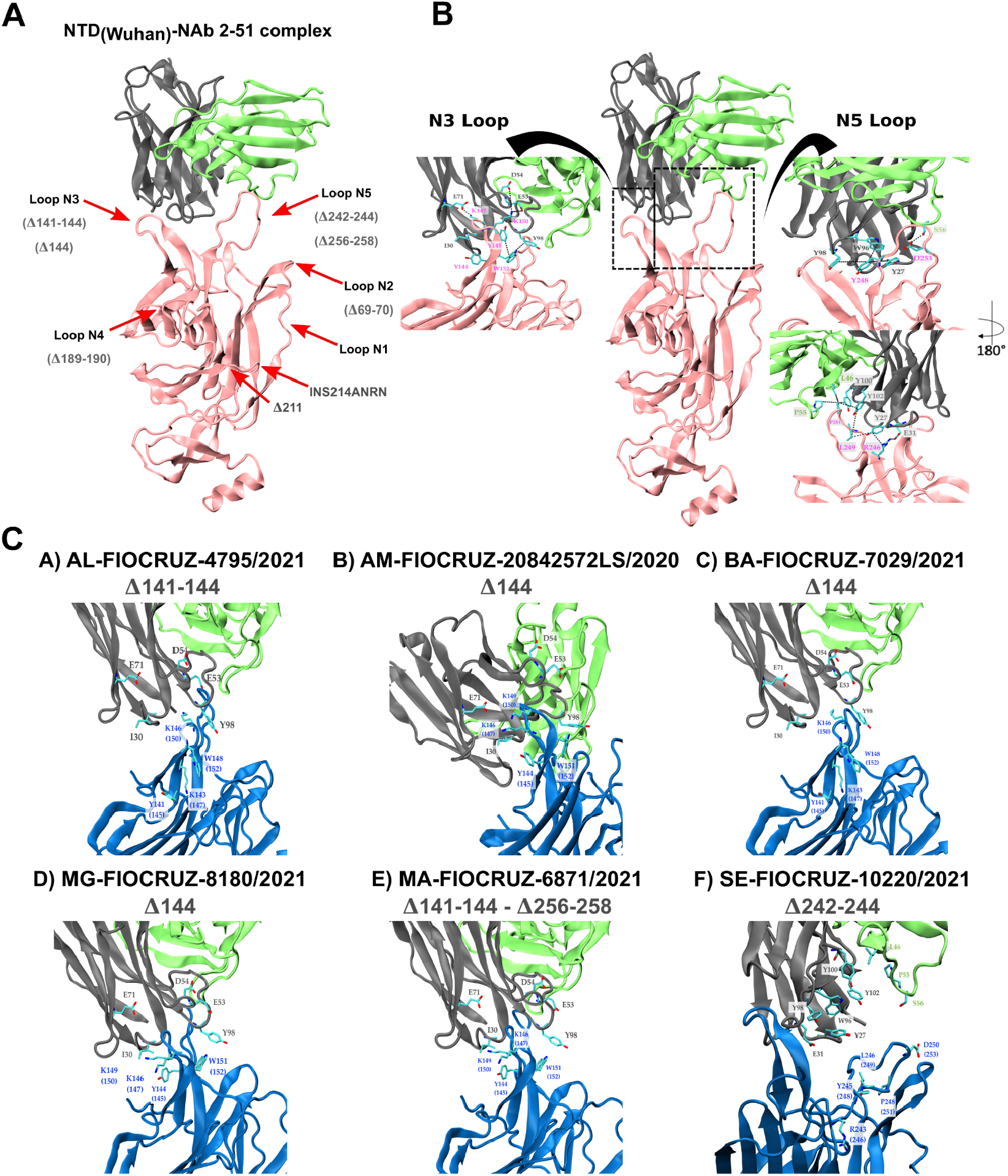
Representation of the Spike NTD 3D structure of wild type (pink) and NTD deleted variants (colored in blue) complexed to the NAb 2-51 heavy (gray) and light (green) chains. **A)** Relative position of the five NTD loops (red arrows) and the NTD deletions detected in our sample. **B)** Native interactions of mAb NAb 2-51 with N3 (left close-up) and N5 (right close-up) loops on the 3D structure of the wild type Spike NTD antigenic supersite. The N5 loop representation is also rotated 180° around its z-axis. **C)** Potential interactions of mAb NAb 2-51 with N3 and N5 loops on the 3D structure of the Spike NTD of N3 and N5 deleted variants. Residues making contact in the interface are depicted in the licorice representation, with carbon atoms in cyan, nitrogen atoms in blue and oxygen atoms in red. The dotted lines indicate the interacting residues-pair.

Our analyses corroborate that deletions at RDR2/IR-3 (Δ144, Δ141-144) and RDR4/IR-5 (Δ242-244, Δ256-258) detected in Brazilian sequences impact the N3 and N5 loops’ size and conformation, disrupting the native contacts and reducing the interacting hydrophobic surface accessible area, mainly due to the loss of the hydrophobic pocket (**Figure 3C**). Indels around the N3/N5 loops resulted in a significant loss of interactions (both electrostatic and hydrophobic) that can dramatically impact the binding free energy, and therefore the binding affinity, between those NTD deletion variants and the NAb 2-51. Variant P.1 Δ242-244 displayed the largest loss of interactions, followed by variants N10, P.1 Δ141-144, P.1 Δ144, P.2 Δ144, and B.1.1.28 Δ144 (**Table 2**). The NTD indels Δ69-70, Δ189-190, Δ211 and ins214ANRN did not affect the NTD antigenic-supersite (**Figures 3A**), but they occur at other loops that comprise putative epitope regions covering residues 64-83, 168/173-188 and 209-216 (**Appendix Table 2**) and leads to conformational changes **(Supplementary Figure S2)** which might affect Ab binding outside the NTD antigenic-supersite. These findings suggest that NTD indels detected here probably abrogate the binding of NAb directed against the antigenic-supersite and other epitopes.

**Table 2.** Impact of indels on the binding between SARS-CoV-2 NTDs and NAb 2-51, expressed as loss of putative interactions.

| Variant | ΔH-bond | ΔSalt-bridge | Δpi-stacking | ΔHydrophobic SASA [$\text{\AA}^2$] | Native Contacts Lost |
| --- | --- | --- | --- | --- | --- |
| B.1.1.28 Δ144 | -2 | -3 | -1 | -1 | K147-E71<br>K150-E53<br>K150-D54<br>Y145-Y98 |
| P.2 Δ144 | -2 | -3 | -1 | -104 | K147-E71<br>K150-E53<br>K150-D54<br>Y145-Y98 |
| P.1 Δ144 | -2 | -3 | -1 | -111 | K147-E71<br>K150-E53<br>K150-D54<br>Y145-Y98 |
| P.1 Δ141-144 | -2 | -3 | -1 | -313 | K147-E71<br>K150-E53<br>K150-D54<br>Y145-Y98 |
| N.10<br>Δ141-144<br>Δ256-258 | -3 | -3 | -1 | -439 | Y147-E71<br>K150-E53<br>K150-D54<br>Y145-Y98<br>D253-S56<br>P251-P55<br>P251-L46<br>P251-Y100 |
| P.1 Δ242-244 | -3 | -1 | -2 | -746 | Y248-Y27<br>Y248-W96<br>Y248-Y98<br>L249-Y27<br>R246-E31<br>R246-Y27<br>D253-S56 |

Several studies of SARS-CoV-2 evolution *in vitro* and *ex vivo* also support that NTD indels here observed in Brazilian SARS-CoV-2 VOC and VOI represent a mechanism of ongoing adaptive evolution to escape from dominant neutralizing antibodies directed against the NTD. *In vitro* co-incubation of SARS-CoV-2 with highly neutralizing plasma form COVID-19 convalescent patient, has revealed an incremental resistance to neutralization followed by the stepwise acquisition of indels at N3/N5 loops ^35^. SARS-CoV-2 challenge in hamsters previously treated with anti-NTD mAbs revealed selection of two escape mutants harboring NTD deletions Δ143-144 and Δ141-144 ^14^. Studies of intra-host SARS-CoV-2 evolution in immuno-compromised hosts revealed the emergence of viral variants with NTD deletions at RDR1 (Δ69-70), RDR2 (Δ144 and Δ141-144) and RDR4 (Δ243-244) following therapy with convalescent plasma ^1,3,4,36,37^. Another study revealed the emergence of several Spike gene mutations, including inframe deletions Δ141-143, Δ141-144, Δ145 and Δ211-212, during persistent SARS-CoV-2 infection in two individuals with partial humoral immunity ^38^. Finally, a recent longitudinal analysis of intra-host SARS-CoV-2 evolution during acute infection in one immunocompetent individual revealed the emergence of virus haplotypes bearing deletions Δ144 and Δ141-144 in the NTD following the development of autologous anti-NTD specific antibodies ^39^.

Recent genomic findings showed a sudden landscape change in SARS-CoV-2 evolution since October 2020, coinciding with the independent emergence of VOCs carrying multiple convergent amino acid replacements at the RBD of the S protein ^40^. One hypothesis is that such a major selection pressure shift on the virus genome is driven by the increasing worldwide human population immunity acquired from natural SARS-CoV-2 infection that might also select for convergent deletions at NTD. Our findings suggest that P.1, P.2 and N.10 variants with NTD indels here detected might have evolved to escape from NAb against NTD and could be even more resistant to neutralization than the parental viruses. Notably, the sequential acquisition of RBD and NTD mutations observed in the VOC P.1 recapitulates the evolution pattern of the VOC B.1.351 that first acquired RBD mutations E484K and N501Y and sometime later the NTD deletion Δ242-244 ^7^. The detection of P.1 genomes with convergent NTD deletions with VOCs B.1.1.7 (Δ69-70, Δ144) and B.1.351 (Δ242-244) bring caution about the specificity of published real-time RT-PCR protocols to distinguish different VOCs in Brazil and also alert against use the failure to detect the S gene (due to mutation Δ69-70) by certain tests, known as S gene target dropout ^41,42^, as a definitive proof of circulation of the VOC B.1.1.7 in Brazil.

In summary, these findings suggest that SARS-CoV-2 VOC and VOI are continuously adapting and evolving in Brazil through acquisition of Spike NTD indels. Some variants like P.1 Δ69-70, P.1 Δ144 and P.1-like ins214ANRN might represent newly emergent VOC/VOI and its communitary dissemination, as well as that of VOI N.10, requires careful monitoring. These findings highlight the urgent need to address the SARS-CoV-2 vaccines’ efficacy towards emergent SARS-CoV-2 variants carrying both RBD and NTD mutations and deletions of concern and the risk of ongoing uncontrolled community transmission of SARS-CoV-2 in Brazil for the generation of more transmissible variants. The recurrent emergence of NTD ins214 variants in different SARS-CoV-2 lineages circulating in the Americas and Europe since November 2020 and its impact on vaccine efficacy also deserves further attention.

## Material and Methods

### SARS-CoV-2 and SARS-CoV-2-related coronavirus (SC2r-CoV) sequences

Our genomic survey of SARS-CoV-2 positive samples sequenced by the Fiocruz COVID-19 Genomic Surveillance Network between 12th March 2020 and 28th February 2021 identified 11 sequences with mutations of concern in the RBD and indels in the NTD (**Appendix Table 1**). The SARS-CoV-2 genomes were recovered using Illumina sequencing protocols as previously described ^43,44^. The FASTQ reads obtained were imported into the CLC Genomics Workbench version 20.0.4 (Qiagen A/S, Denmark), trimmed, and mapped against the reference sequence EPI_ISL_402124 available in EpiCoV database in the GISAID (https://www.gisaid.org/). The alignment was refined using the InDels and Structural Variants module. Additionally, the same reads were imported in a different pipeline ^45^ based on Bowtie2 and bcftools ^46^ mapping and consensus generation allowing us to further confirm the indels supported by paired-end reads, removing putative indels with less than 10x of sequencing depth and with mapping read quality score below to 10 for all samples sequenced in this study. BAM files were used as input to generate sequencing coverage plots onto indels using the Karyoploter R package ^47^. Sequences were combined with SARS-CoV-2 and SC2r-CoV from bats and pangolins available in the EpiCoV database in GISAID by 1st March 2021 (**Appendix Table 1**). This study was approved by the FIOCRUZ-IOC (68118417.6.0000.5248 and CAAE 32333120.4.0000.5190) and the Amazonas State University Ethics Committee (CAAE: 25430719.6.0000.5016) and the Brazilian Ministry of the Environment (MMA) A1767C3.

### Maximum Likelihood Phylogenetic Analyses

SARS-COV-2 sequences here obtained were aligned with high quality (<1% of N) and complete (>29 kb) lineages B.1.1.28, P.1, P2 and B.1.1.33 sequences that were available in EpiCoV database in the GISAID (https://www.gisaid.org/) at March 1st, 2021 and subjected to maximum-likelihood (ML) phylogenetic analysis using IQ-TREE v2.1.2 ^48^. The S amino acid sequences from selected SARS-CoV-2 and SC2r-CoV lineages available in the EpiCoV database were also aligned using Clustal W ^49^ adjusted by visual inspection.

### Structural Modeling

The resolved crystallographic structure of SARS-CoV-2 NTD protein bound to the neutralizing antibody 2-51 was retrieved from the Protein Databank (PDB) under the accession code 7L2C ^33^. Missing residues of the chain A, corresponding to the NTD coordinates, were modeled using the user template mode of the Swiss-Model webserver (https://swissmodel.expasy.org/) ^50^ and was used as starting structure for the NTD wildtype. This structure was then used as a template to model the NTD variants using the Swiss-Model webserver. The modeled structures of the NTDs variants were superimposed onto the coordinates of the PDB ID 7L2C to visualize the differences between the NTD-antibody binding interfaces. Image rendering was carried out using Visual Molecular Dynamics (VMD) software ^51^. The NTD-antibody complexes were geometry optimized using a maximum of 5,000 steps or until it reached a convergence value of 0.001 REU (Rosetta energy units) using the limited-memory BroydenFletcher-Goldfarb-Shanno algorithm, complying with the Armijo-Goldstein condition, as implemented in the Rosetta suite of software 3.12 ^52^. Geometry optimization was accomplished through the atomistic Rosetta energy function 2015 (REF15), while preserving backbone torsion angles. Protein-protein interface analyses were performed using the Protein Interactions Calculator (PIC) webserver (http://pic.mbu.iisc.ernet.in/)^53^, the ‘Protein interfaces, surfaces and assemblies’ service (PISA) at the European Bioinformatics Institute (https://www.ebi.ac.uk/pdbe/pisa/pistart.html) ^54^ and the InterfaceAnalyzer protocol of the Rosetta package interfaced with the RosettaScripts scripting language ^55^. For the interfaceAnalyzer, the maximum SASA that is allowed for an atom to be defined as buried is 0.01 Å^2^, with a SASA probe radius of 1.2 Å.

### Epitope prediction

Epitopes in the NTD region were predicted by the ElliPro Antibody Epitope Prediction server ^56^. NTD are shown as predicted linear epitopes when using PDB accession codes 6VXX ^57^ and 6VSB ^58^, (structural coordinates corresponding to the entire S protein), along with a minimum score of 0.9, *i*.*e*., a highly strict criterion.

## Data Availability

All genome sequences used in this study were deposited in GISAID and corresponding IDs can be found in the manuscript.

## Acknowledgements

The authors wish to thank all the health care workers and scientists who have worked hard to deal with this pandemic threat, the GISAID team, and all the EpiCoV database’s submitters, GISAID acknowledgment table containing sequences used in this study are attached to this post (**Appendix Table 3**). We also appreciate the support of the Fiocruz COVID-19 Genomic Surveillance Network (http://www.genomahcov.fiocruz.br/) members, the Respiratory Viruses Genomic Surveillance Network of the General Laboratory Coordination (CGLab), Brazilian Ministry of Health (MoH), Brazilian Central Laboratory States (LACENs) and the Amazonas surveillance teams for the partnership in the viral surveillance in Brazil. Financial support was provided by FAPEAM (PCTI-EmergeSaude/AM call 005/2020 and Rede Genômica de Vigilancia em Saúde - REGESAM); Ministério da Ciência, Tecnologia, Inovações e Comunicações/Conselho Nacional de Desenvolvimento Científico e Tecnológico - CNPq/Ministério da Saúde - MS/FNDCT/SCTIE/Decit (grants 402457/2020-9 and 403276/2020-9); Inova Fiocruz/Fundação Oswaldo Cruz (Grants VPPCB-007-FIO-18-2-30 and VPPCB-005-FIO-20-2-87) and INCT-FCx (465259/2014-6). Computer allocation was partly granted by the Brazilian National Scientific Computing Center (LNCC). FGN, GLW, RDL and GB are supported by the CNPq through their productivity research fellowships (306146/2017-7, 303902/2019-1, 425997/2018-9 and 302317/2017-1 respectively). G.B. is also funded by the Fundação Carlos Chagas Filho de Amparo à Pesquisa do Estado do Rio de Janeiro – FAPERJ (Grant number E-26/202.896/2018).

