## Appendix Table 1 for "The ongoing evolution of variants of concern and interest of SARS-CoV-2 in Brazil revealed by convergent indels in the amino (N)-terminal domain of the Spike protein"

**Appendix Table 1.** SARS-CoV-2 sequences available at EpiCoV database in the GISAID (https://www.gisaid.org/) that displayed an insert motif in positions 214-216 of the Spike protein.

| **Lineage** | **Number** | **Motif** | **Sampling dates** |
| --- | --- | --- | --- |
| A.2.4 | 52 | AAG | 05/12/2020-01/03/2021 |
| B | 1 | AKKN | 05/03/2020 |
| B | 2 | KRI | 28/12/2020-25/01/2021 |
| B.1 | 7 | TDR | 17/12/2020-16/02/2021 |
| B.1.1.7 | 1 | KFH | 12/02/2021 |
| B.1.177 | 1 | KLGP | 13/11/2020 |
| B.1.2 | 1 | DLA | 31/01/2021 |
| B.1.214 | 80 | TDR | 05/12/2020-22/02/2021 |
| B.1.429 | 1 | AQER | 15/01/2021 |
