## Appendix Table 2 for "The ongoing evolution of variants of concern and interest of SARS-CoV-2 in Brazil revealed by convergent indels in the amino (N)-terminal domain of the Spike protein"

**Appendix Table 2.** SARS-CoV-2 epitopes predicted by the ElliPro Antibody Epitope Prediction server in the NTD domain of the Spike rpotein.

|  | **Start** | **End** | **Peptide** | **Number of**  **Residues** | **Score** | **IR** | **RDR** |
| --- | --- | --- | --- | --- | --- | --- | --- |
| **6VXX** | 64 | 83 | WFHAIHDNPV | 10 | 0.957 | IR-2 | RDR1 - 𝚫60-75 |
|  | 94 | 103 | STEKSNIIRG | 10 | 0.956 | - | - |
|  | 109 | 114 | TLDSKT | 6 | 0.908 | - | - |
|  | 118 | 130 | LIVNNATNVVIKV | 13 | 0.924 | - | - |
|  | 132 | 165 | EFQFCNDPFLGVN | 13 | 0.964 | IR-3 | RDR2 - 𝚫139-146 |
|  | 168 | 188 | FEYVSFKN | 8 | 0.919 | - | - |
|  | 209 | 216 | PINLVRDL | 8 | 0.933 | IR-4 | RDR3 - 𝚫210-212 |
|  | 239 | 265 | QTLLALHAAY | 10 | 0.958 | IR-5 | RDR4 - 𝚫242-248 |
| **6VSB** | 63 | 83 | TWFHDNPV | 8 | 0.932 | IR-2 | RDR1 - 𝚫60-75 |
|  | 94 | 103 | STNIIRG | 7 | 0.949 | - | - |
|  | 110 | 113 | LDSK | 4 | 0.903 | - | - |
|  | 120 | 127 | VNNATNVV | 8 | 0.938 | - | - |
|  | 132 | 165 | EFQFCNDPFFRVYSSANN | 18 | 0.959 | IR-3 | RDR2 - 𝚫139-146 |
|  | 173 | 188 | QPFLKN | 6 | 0.921 | - | - |
|  | 209 | 216 | PINLVRDL | 8 | 0.928 | IR-4 | RDR3 - 𝚫210-212 |
|  | 240 | 265 | TLLALHGAAAY | 11 | 0.969 | IR-5 | RDR4 - 𝚫242-248 |
