## Supplementary figures and images for "The ongoing evolution of variants of concern and interest of SARS-CoV-2 in Brazil revealed by convergent indels in the amino (N)-terminal domain of the Spike protein"

### Supplementary Figure S1

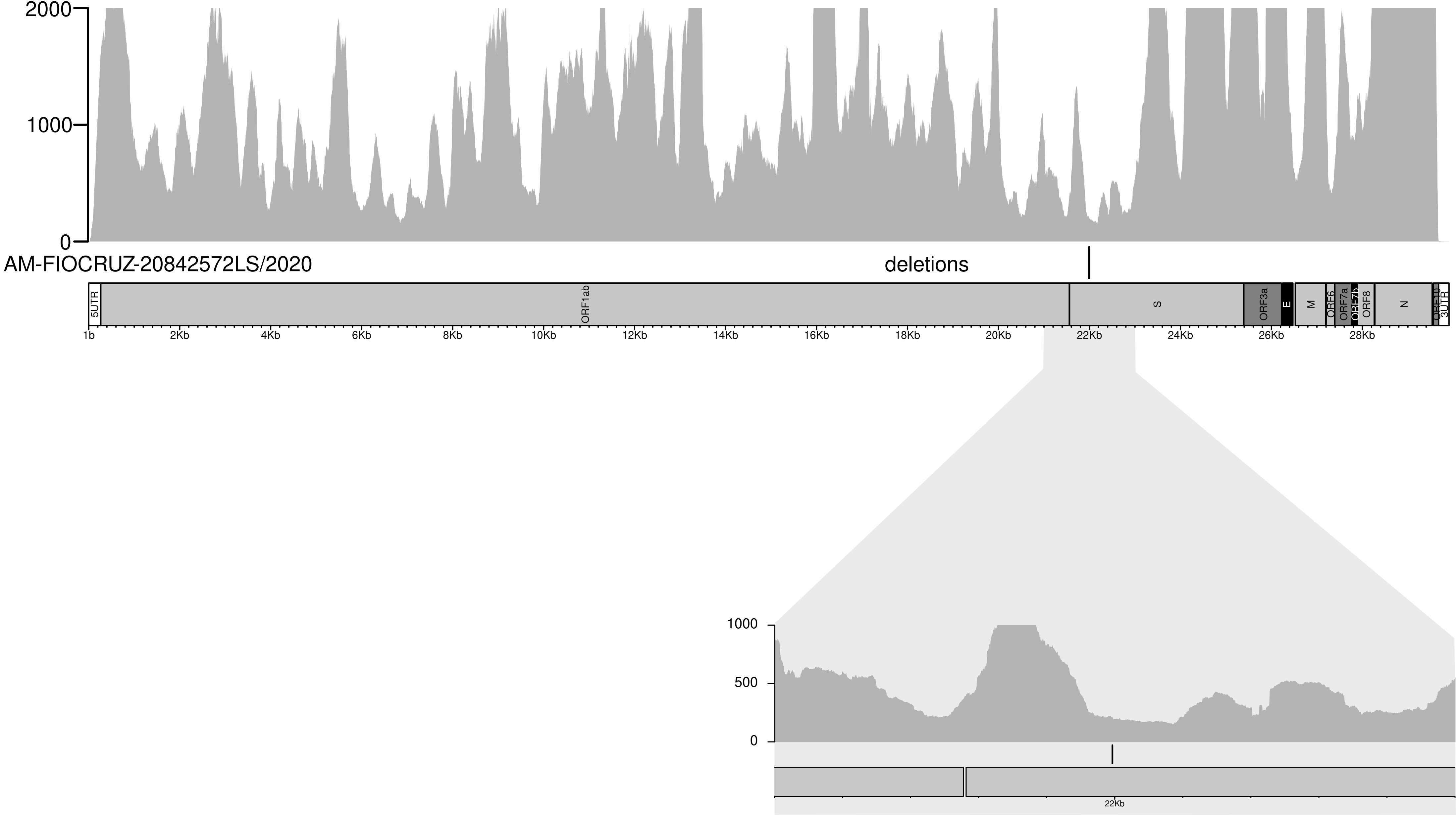

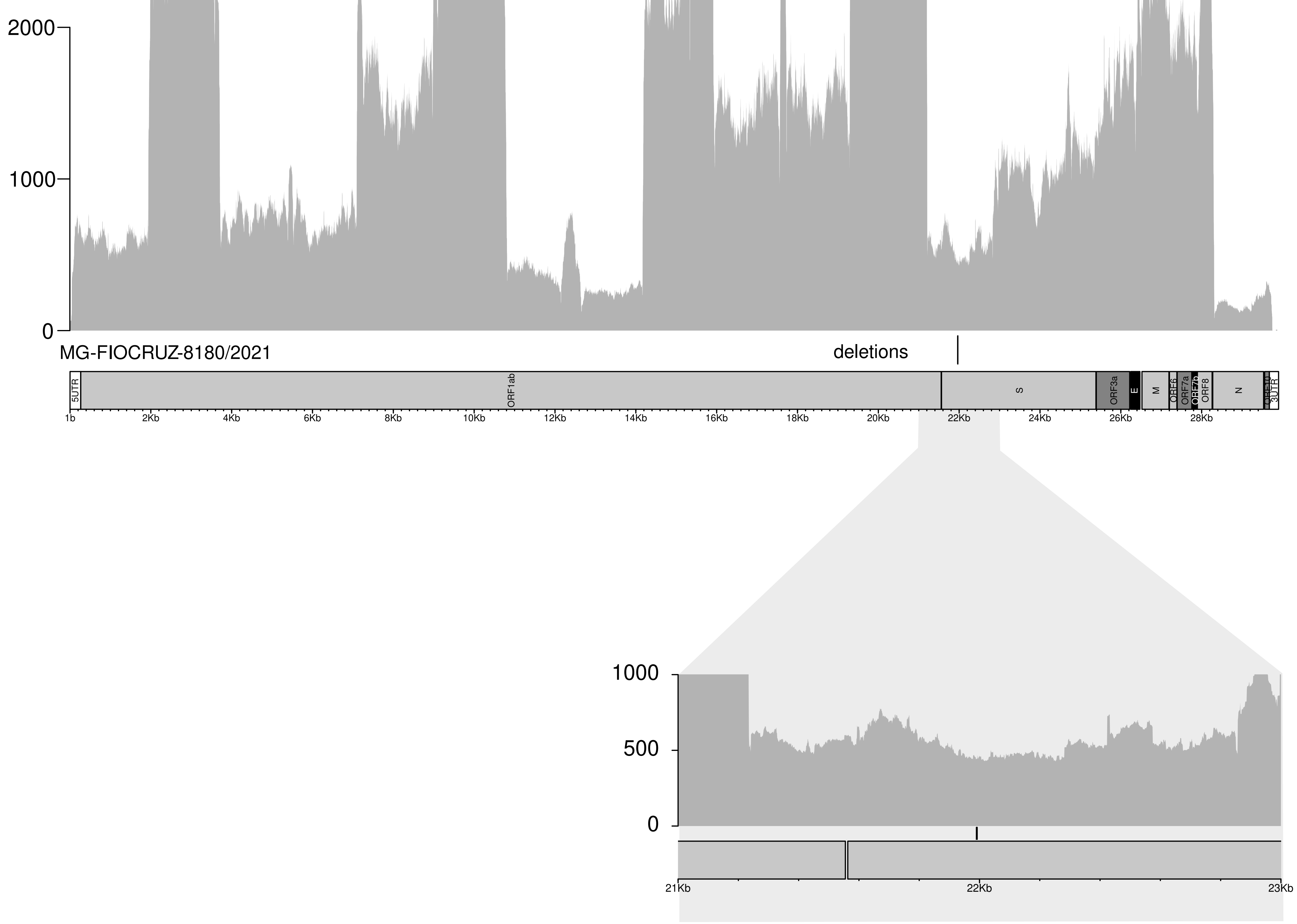

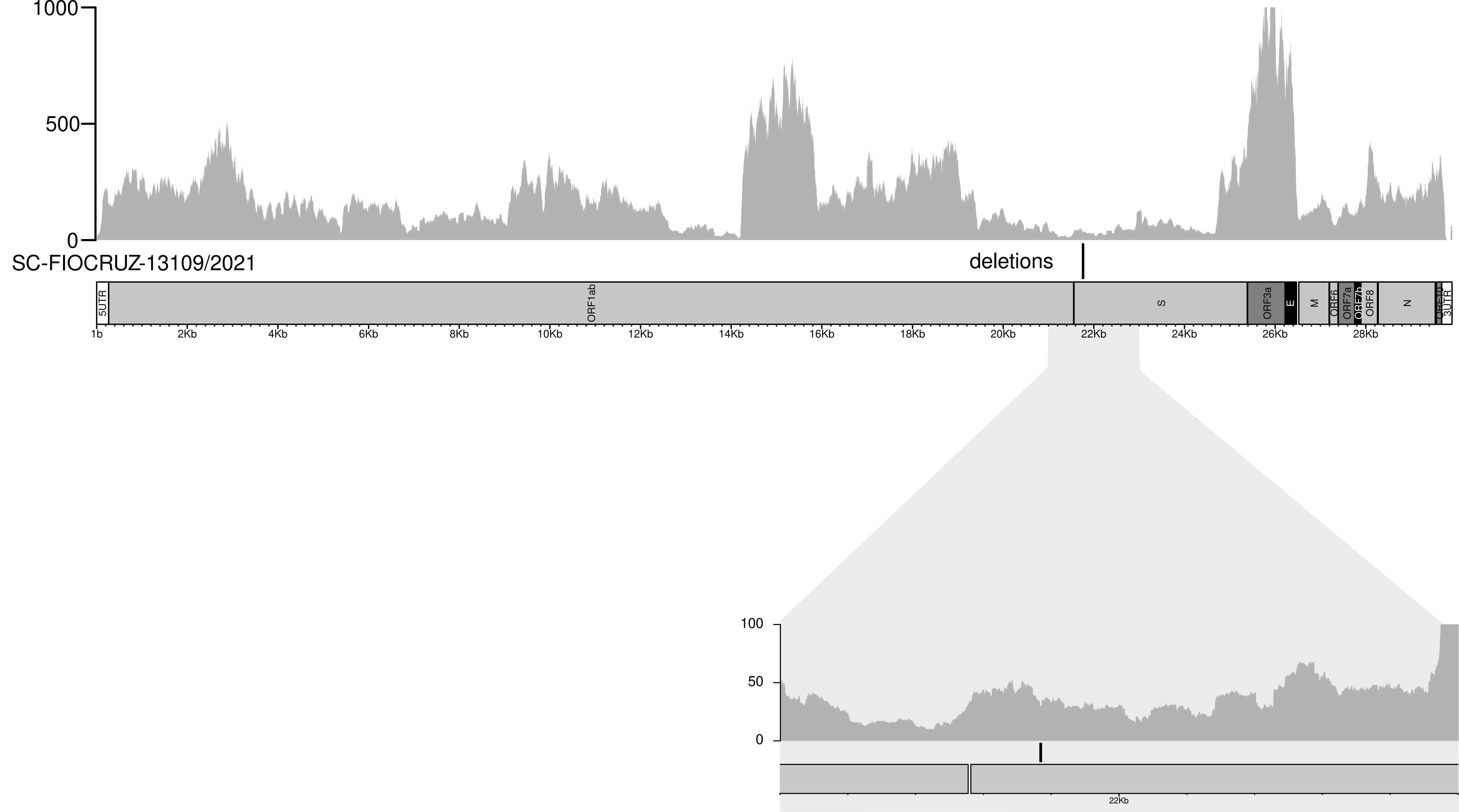

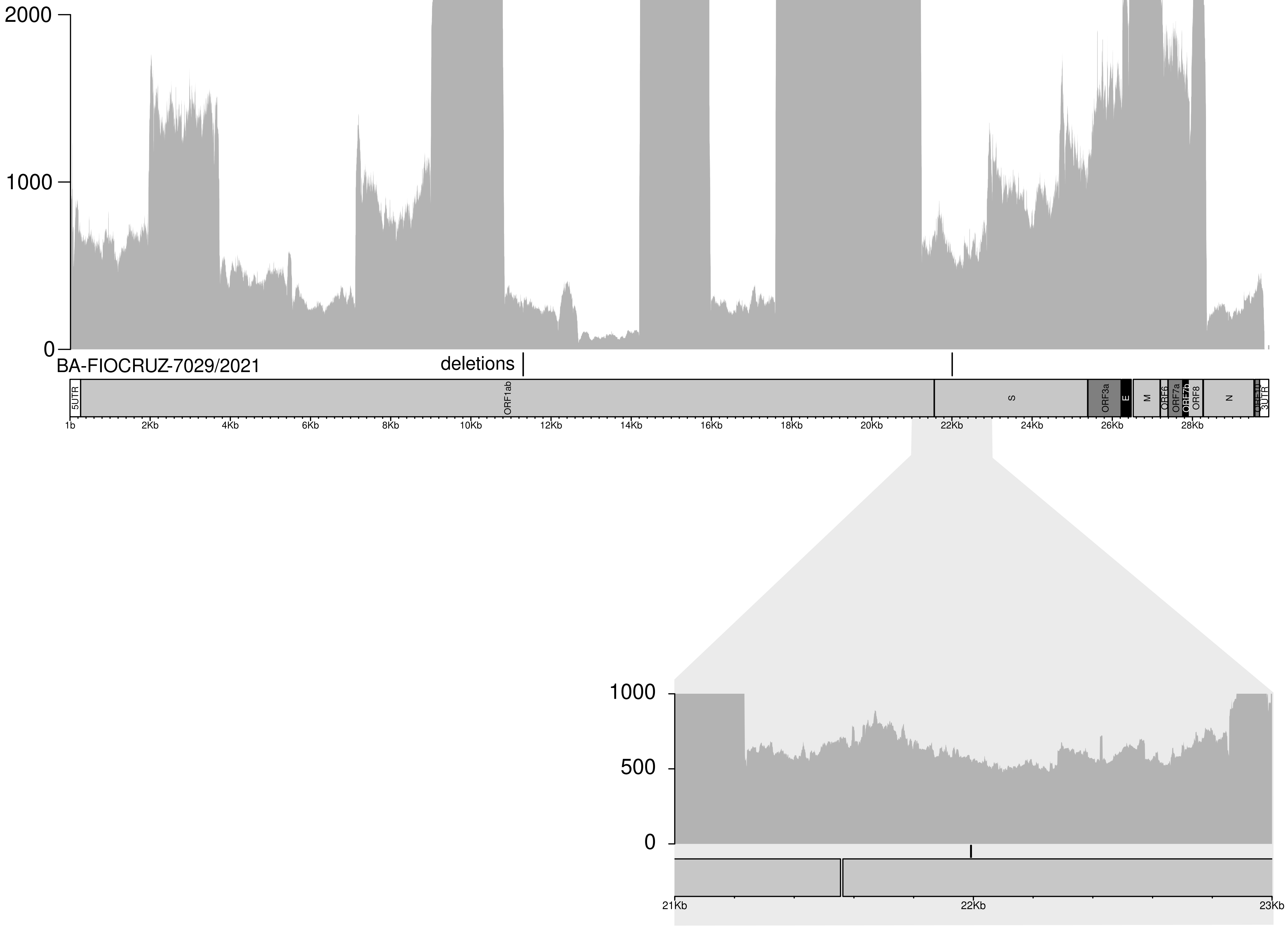

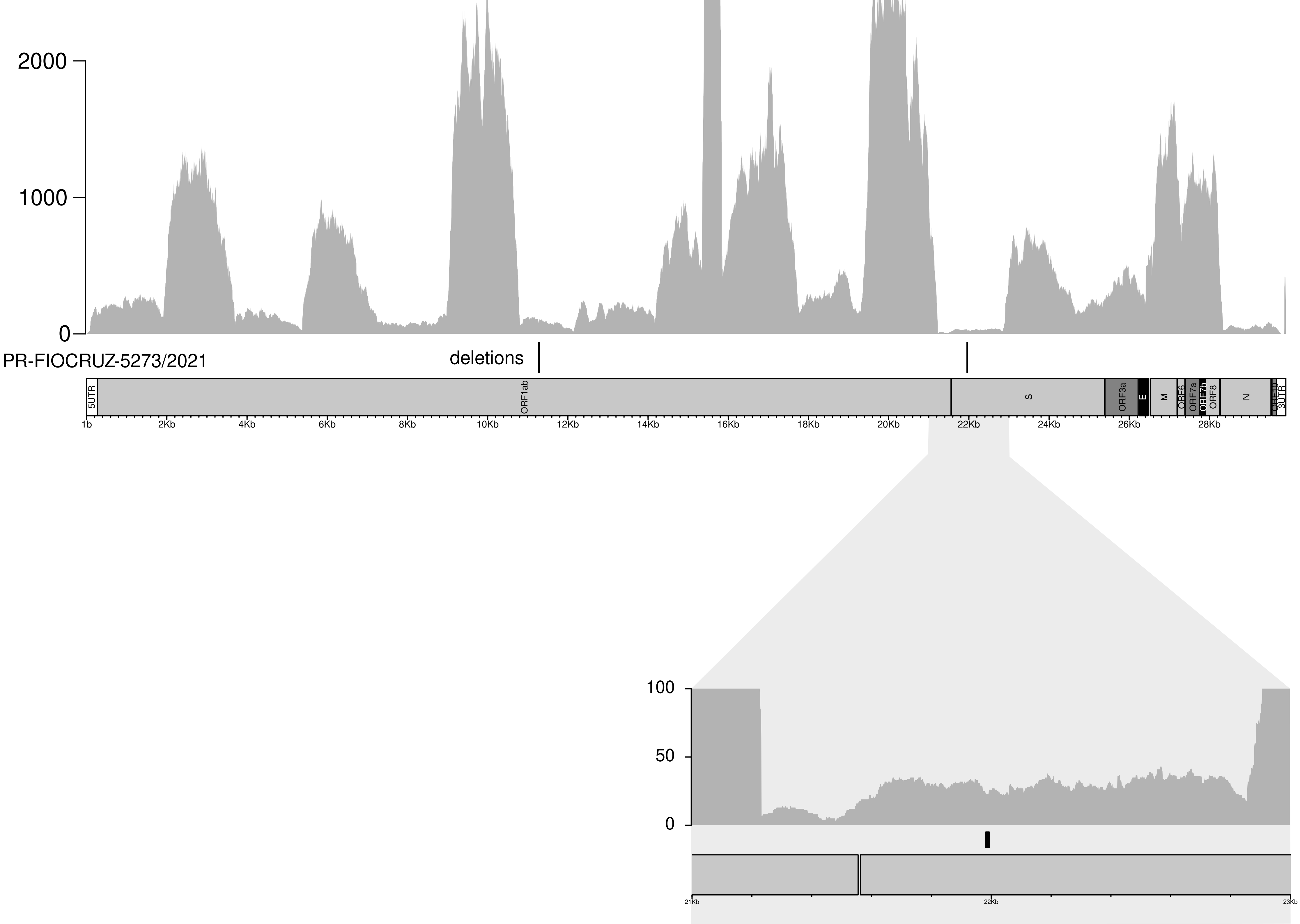

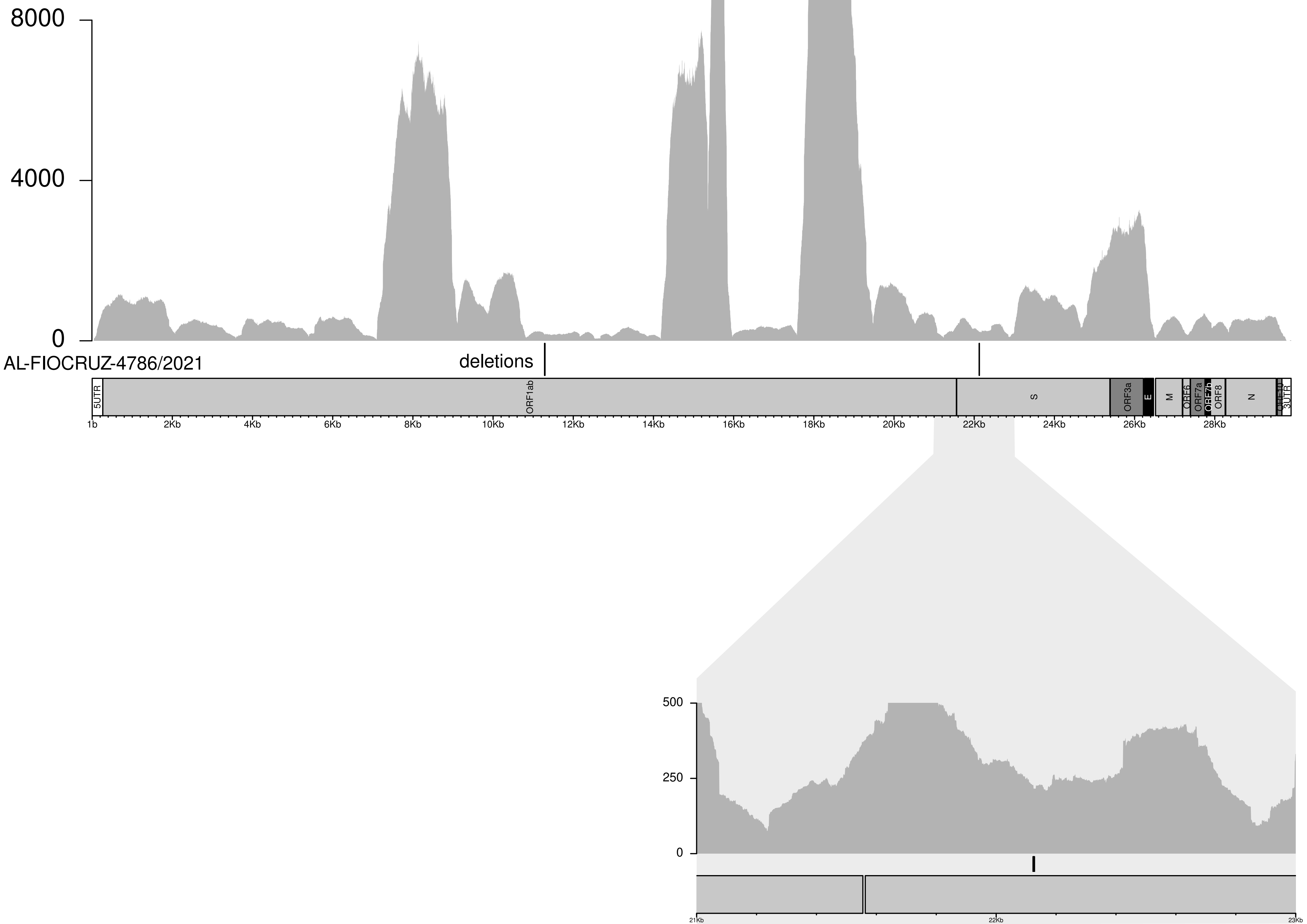

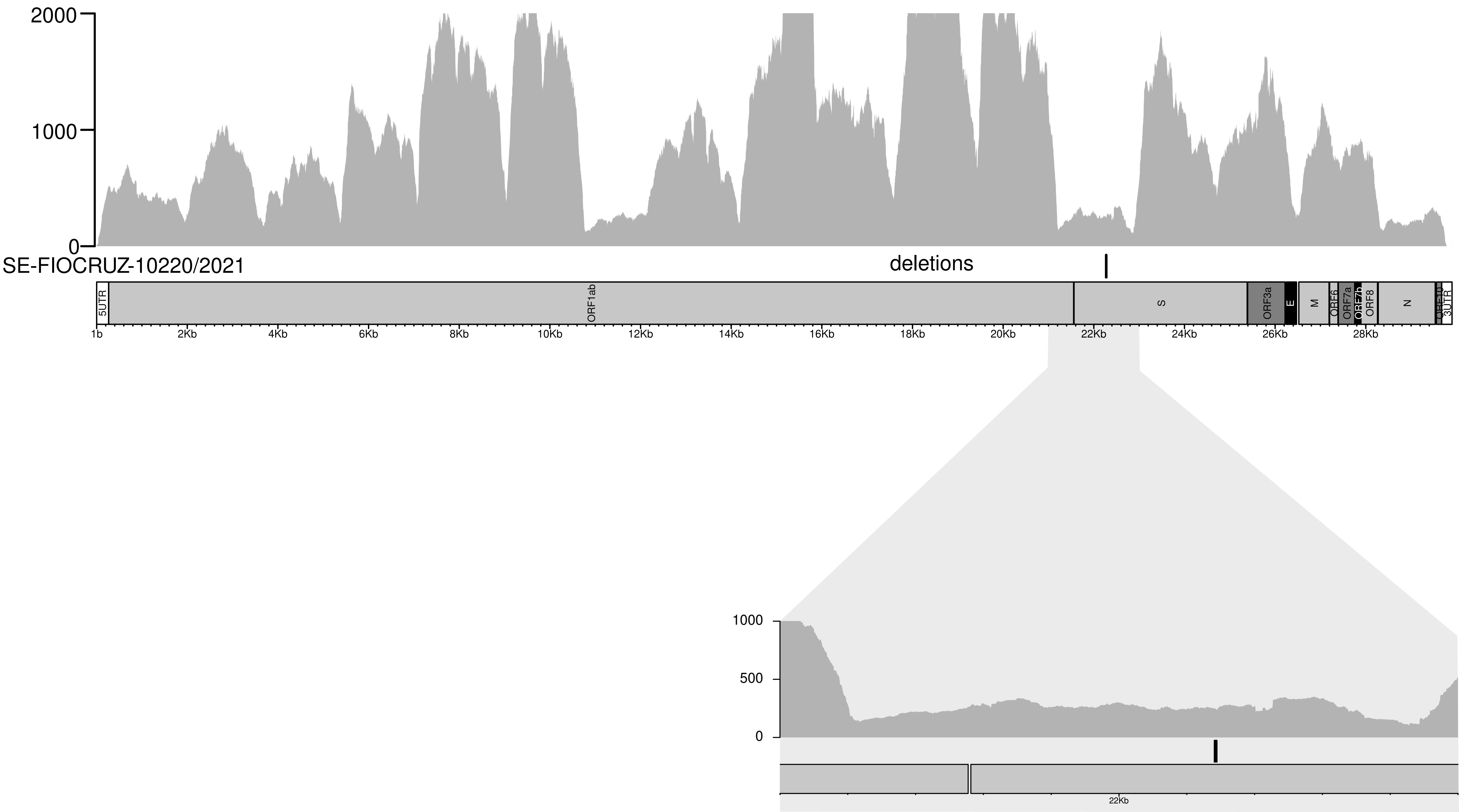

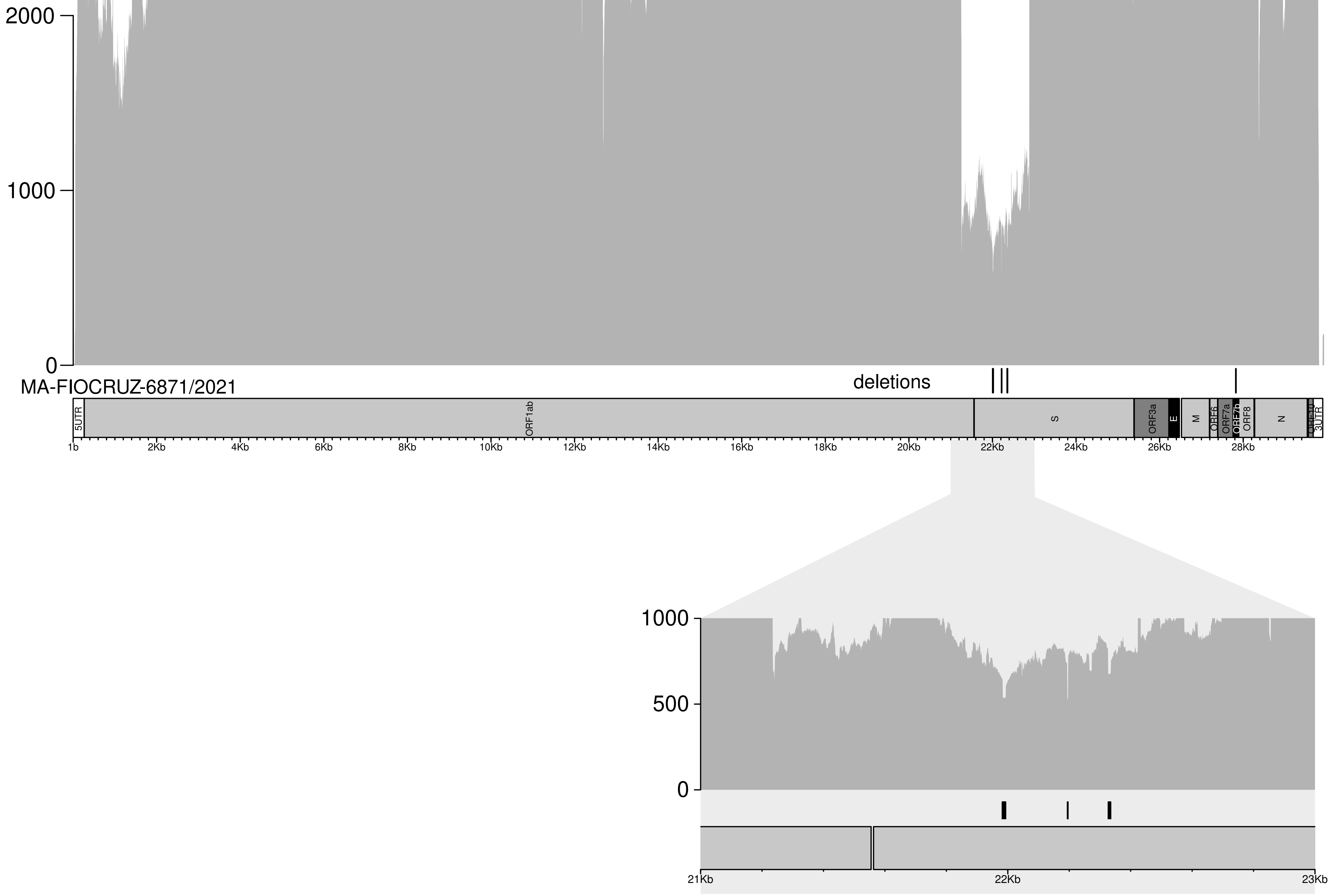

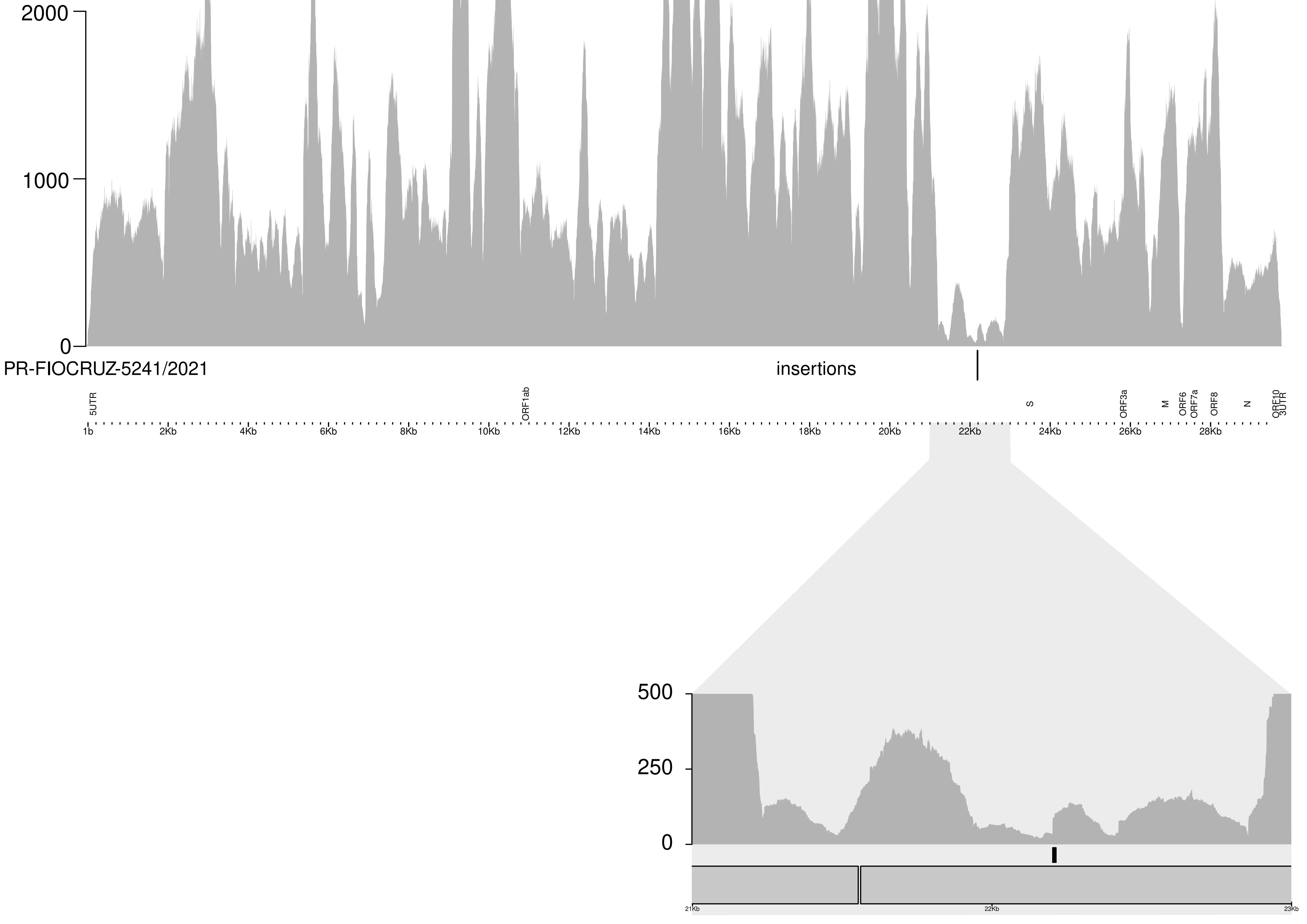

### Supplementary Figure S2

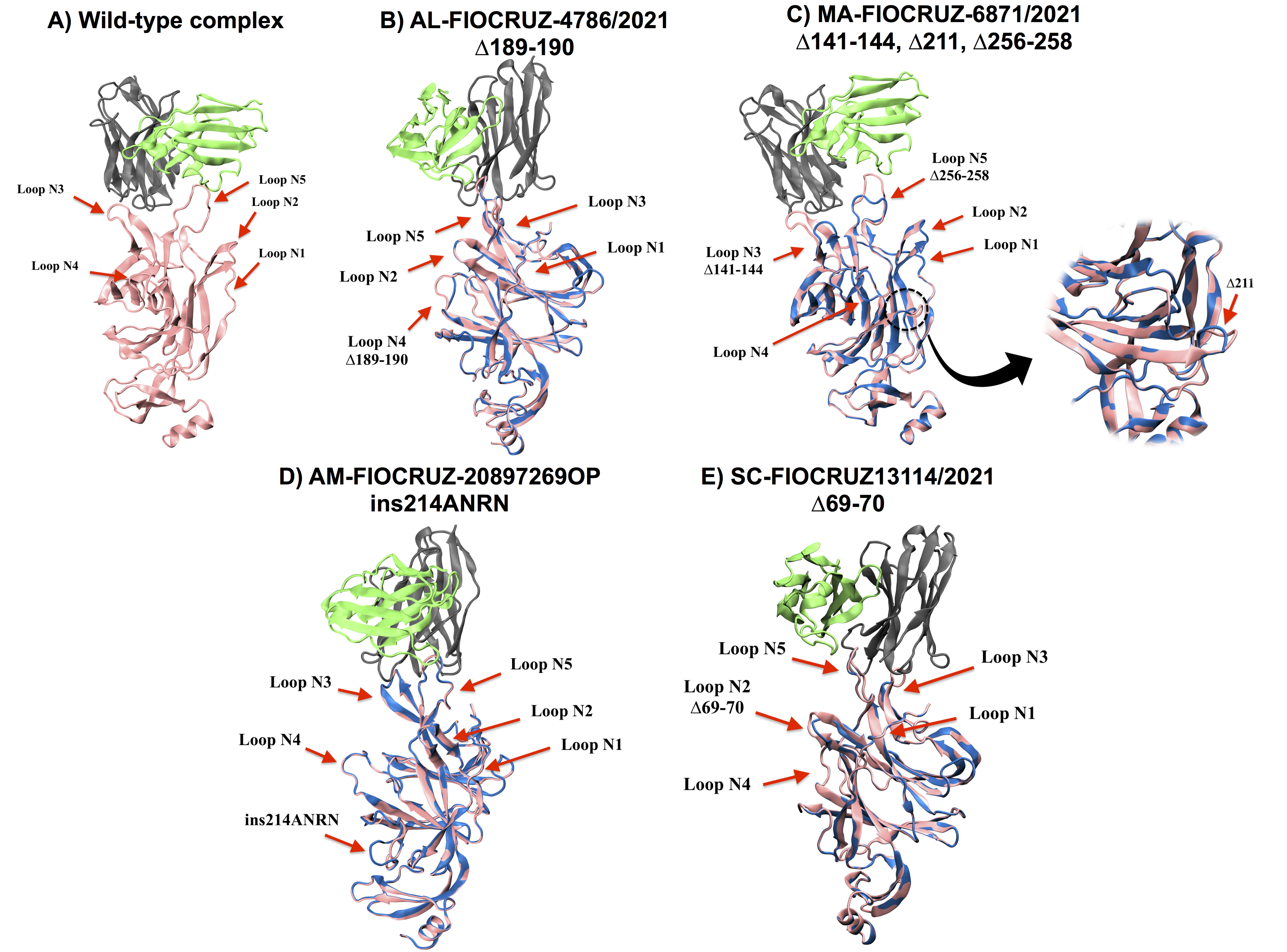
